# Care Phenotypes In Critical Care

**DOI:** 10.1101/2025.01.24.25320468

**Authors:** LL. Weishaupt, T. Wang, J. Schamroth, P. Morandini, J. Matos, LM Hampton, J. Gallifant, A. Fiske, N. Dundas, K. David, LA. Celi, A. Carrel, J. Byers, G. Angelotti

**Affiliations:** Harvard & MIT, Health Sciences and Technology, Cambridge, MA, United States; Ronald Reagan UCLA Medical Center, Los Angeles, United States; Faculty of Population Health Sciences, UCL, London, UK; IRCCS Humanitas Research Hospital, Artificial Intelligence Center, Milan, Italy; Nuffield Department of Orthopaedics, Rheumatology and Musculoskeletal Sciences, University of Oxford, UK; Massachusetts Institute of Technology, Electrical Engineering and Computer Science, Cambridge, MA, United States; Massachusetts Institute of Technology, Laboratory for Computational Physiology, Cambridge, MA, United States; Institute of History and Ethics in Medicine, Department of Preclinical Medicine, TUM School of Medicine and Health, Technical University of Munich; UC Berkeley Department of Bioengineering, CA, United States; Beth Israel Deaconess Medical Center, Respiratory Therapy, Boston, MA, United States; Imperial College London, London, UK; Dalle Molle Institute for Artificial Intelligence, USI-SUPSI, Lugano, Switzerland

## Abstract

The Social Determinants of Health (SDoH) have long been recognised as significant drivers of health inequalities. Within healthcare settings, large EHR datasets have increasingly enabled the use of machine learning (ML) to explore how patient background and demographic factors mediate and predict clinical outcomes. The intensive care unit (ICU) in particular provides a rich source of data for such research. However, major limitations with current approaches persist, including (i) overreliance on individual demographic labels or measures of difference, (ii) the impracticality of highly intersectional patient groups and (iii) that the underlying accuracy and validity of these demographic constructs is low.

The main objective of this study was to take a novel approach, to first understand who within the ICU setting receives sub-standard care and use this to create new, objective labels based on quality of care and outcomes (‘Care Phenotypes’) when different patients interface with the health system. Using the MIMIC-IV database, we focused on highly protocolised, essential care procedures (turning, mouth care) in mechanically ventilated ICU patients. We performed a series of regression analyses to understand in which patients treatment deviated from these protocols.

In a cohort of 8,919 ICU patients undergoing IMV, consistent patterns in sup-optimal protocol adherence for certain groups, notably heavier patients. Compared to equivalently sick peers, for every extra weight decile, a patient can expect a reduction of one percentile in frequency of turning care (0.0760 turning interval percentile per weight percentile, p<0.05). Furthermore, patients who receive fewer turnings should also expect to receive fewer mouth care procedures, in a quantile ratio of 1 to 5 (0.2055 mouth care interval quantile per turning interval quantile, p<0.05).

The findings in this initial analysis provide support to the concept of first looking at the actual care delivered to patients to illuminate the relationship between patient demographics and outcomes of interest. The ‘Care Phenotypes’ approach has the potential to improve fairness evaluations for machine learning in healthcare, support causal inference research and play a larger role in research into healthcare disparities.

## INTRODUCTION

### Social determinants of health in the era of machine learning

The Social Determinants of Health (SDoH) have long been recognised as significant drivers of health inequalities. The SDoH include socioeconomic status, sex and gender, ethnic background and education levels, all of which can have significant impacts on health outcomes.[1] Adverse structural and social factors including poverty, racism and discrimination by gender, ethnicity or class can also be understood as adverse social determinants of health that are important drivers of health outcomes.[2]

One lens for understanding how these demographic factors lead to inequities in health outcomes is to explore how they manifest within a healthcare setting. An advantage of many healthcare settings (albeit largely those in rich, Western nations) is that electronic health records (EHRs) and associated data collection provides a rich source of information for exploring the associations between a patients’ background, environment, disease and health outcomes. The intensive care unit (ICU) in particular is, at least in theory, an optimal setting for research exploring disparities in care delivery and outcomes across different patient demographics.[3] ICU care typically involves invasive treatment with continuous monitoring of physiological status through medical equipment, generating vast amounts of granular clinical data relating to all aspects of care, much of it captured within EHR.

As the use of machine learning (ML) applied to biomedical and health systems research has expanded, researchers have adopted these tools to help probe the relationship between background SDoH factors and care outcomes within the ICU [24]. ML offers potentially powerful tools for eliciting relationships and predicting outcomes by iteratively training models on ever greater volumes of clinical data.

### The limitations of demographic labels

To date, there has been no shortage of research into how and why disparities manifest within the ICU. Typically, research has explored differences in key health outcomes as well as differences in the type, or amount, of care ICU patients receive, according to different demographic factors. A wide range of prospective and retrospective studies have explored the relationship between ICU care, health outcomes and i. patient race or ethnicity, [4][5][6][7][8][9] ii. patient sex or gender, [10][11][12][13][14] and iii. socioeconomic status [15] [16][17]. Furthermore, there has increasingly been recognition that the way data is collected within the ICU clinical setting is itself inherently prone to bias along demographic lines which can impact outcomes. One notable example is that patients with darker skin pigmentation are at risk of real-world harm through lower oxygen levels that go undetected due to poorly calibrated pulse-oximeter devices producing inaccurate Sp02 readings. [18] Similar calibration issues have been identified in other ICU medical devices, which risks encoding bias into clinical data. This in turn becomes more problematic when the large EHR datasets generated from routine ICU data are used to train machine learning models.

Despite progress in this area in recent years, it is apparent from the literature that the use of existing demographic categories for decades has not meaningfully progressed our understanding of health disparities nor facilitated the development of effective tools for mitigating them. The current use of demographic labels for SDoH research within the ICU has major limitations, which are brought into focus when we consider the data required for training machine learning algorithms:

i. Currently, we rely too heavily on existing metadata categories. Investigating discrepancies in care or outcomes only by individual measures of difference (e.g. race) are too blunt, arbitrary and insufficient for exploring complex, subtle and intersectional questions of disparities that can occur across numerous axes (race, sex, gender, weight, income level, geography). [25,26,27,31] There is mounting evidence that it is the intersections of these factors that will provide the real insights.
ii. Furthermore, attempts at intersectional analyses considering numerous demographic factors also pose major challenges, slicing this pie more finely results in smaller sample sizes which are cumbersome, impractical and often inadequate for research purposes.
iii. Finally, a more fundamental problem exists at the heart of such research efforts, which is that the underlying accuracy and validity of these demographic constructs captured within EHR data may be low. Inaccurate or incomplete data capture about patient background is rife throughout EHR data, posing major issues in data quality. [26, 27,31] The resulting labels relating to patient ethnicity, gender and class may lack fidelity, which can seriously undermine utility and application of this data in health systems research, including for machine learning.

### Care Phenotypes: A new approach

Here, we take a novel approach for exploring how differences in ICU care link to individual patient backgrounds. Our aim was to start by understanding who within the ICU setting receives sub-standard care, and then use ML techniques to provide new, objective labels based on quality of care and outcomes (‘Care Phenotypes’) when different patients interface with the health system. Care phenotypes are based on the sub-type of care patients actually receive. Recognising that we cannot entirely ignore the use of demographic labels in health services research, in this paper we outline how research can move from a demographic-first approach to an outcomes and care quality-first approach.

Within the ICU, strict guidelines are designed to standardize and optimize patient care, ensuring that every patient receives high quality, evidence-based care. The effectiveness of these guidelines hinges not just on their scientific validity but also on their uniform application across diverse patient groups. Adherence to guidelines should, in principle, be equitable across all patients. In practice, clinical factors, human systems factors as well socioeconomic factors, ethnic background or the healthcare provider’s implicit biases can lead to variations in adherence to these guidelines, with some receiving less or lower quality care than others. [4,28,29,30] Focusing on adherence to specific ICU protocols therefore provides a valuable yardstick to scrutinise care quality and how an individual patient’s care was actually delivered.

Invasive Mechanical Ventilation (IMV) is a critical intervention in ICU for patients experiencing severe respiratory failure. While life-saving, IMV necessitates rigorous care protocols, including regular patient repositioning (turning frequency) and comprehensive oral care (mouth care) to mitigate complications, such as pressure ulcers and ventilator associated pneumonia respectively. Inconsistent adherence to these protocols can result in adverse outcomes, underscoring the importance of examining care consistency across different patient groups.[19] From a data analysis perspective, protocol adherence vis-a-vis common and repetitive tasks such as turning provide valuable, granular data for analysis, yet remain highly clinically salient, because if overlooked, low adherence to the turning protocol can have serious effects.

These routine procedures (turning, mouth care) were also chosen because they are clear and unambiguous to derive from EHRs. High degrees of certainty regarding treatment timings and magnitude in the dataset allows for minimization of the effect of unobserved confounders, providing some reassurance that results obtained were due to the variable under investigation rather than another unadjusted behavior in how the treatment was recorded.

Through discussion amongst an interdisciplinary team of ICU clinicians, machine learning experts and health service researchers, hypotheses were established regarding which demographic factors may be of greatest relevance given our chosen measure of turning. The team chose to focus on patient weight (kg) for two main reasons. Firstly, the logic of a possible causal link between weight and the policy under investigation (turnings) is well defined from an operational standpoint and supported by clinical experience in discussions with nursing and other clinical staff. Secondly, patient weight is a common predictor used in ML model development to characterize patients and has advantages as a continuous variable that allows for stratification and grouping.

## METHODS

### Data Sources

The MIMIC-IV database has been used as the primary data source for this study. The MIMIC-IV is a large database containing de-identified ICU electronic health record (EHR) data collected between 2008 and 2019 from the Beth Israel Deaconess Medical Center in Boston, MA. MIMIC contains demographic profiles of admitted patients, such as self-identified race and ethnicity, English fluency, and insurance. These features allow us to investigate whether disparities linked to weight are stable across diverse sub-populations. Similarly, although de-identified and aggregated, MIMIC retains partial temporal traceability, allowing us to examine whether weight disparities evolve as clinical protocols change. MIMIC-IV contains data for over 65,000 patients admitted to an ICU and over 200,000 patients admitted to the emergency department.[20]

### Inclusion and Exclusion Criteria

We included all patients who underwent Invasive Mechanical Ventilation. We excluded all patients under IMV for less than 24 hours, with missing weights and with weights falling outside the 10-250Kg range, seeFigure 1.

**Figure 1.**
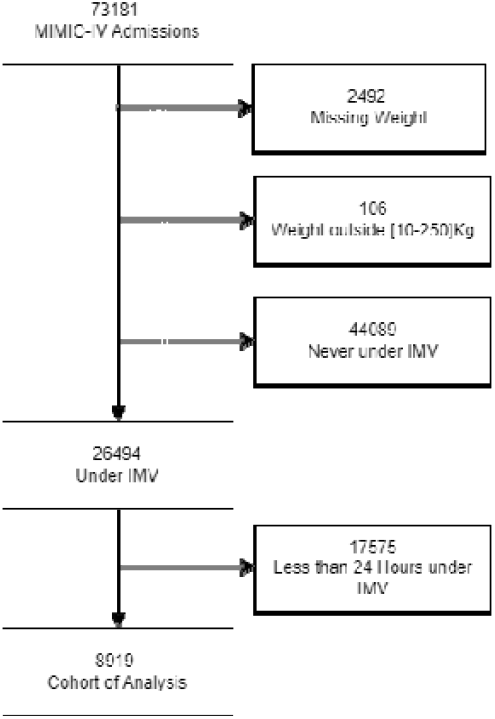
Flow Diagram

## RESULTS

### Main Results

We investigated volumes of turnings and mouth care in a cohort of 8,919 Intensive Care Unit (ICU) patients undergoing Invasive Mechanical Ventilation (IMV) into the MIMIC-IV database. Each patient was characterized by disease severity [14], comorbidities [15] as well as various non-clinical demographic factors, such as ethnicity, type of health insurance and first spoken language. We further characterized patients by their period of hospital admission (from 2008-2019) to track possible clinical policy changes across the years, hospital admission location and type of intensive care unit [4]. Volumes of turnings and mouth care were characterized via average intervals between consecutive administrations. Patients undergoing IMV should be turned regularly, regardless of circumstances, in order to prevent serious consequences [23]. For IMV patients in this MIMIC-IV cohort, ICU protocols for turning and mouth care required/recommended turning every 2 hours and mouth care every 2 to 3 hours. These policies remained in place throughout the 2008-2019 period.

**Table 1A.**
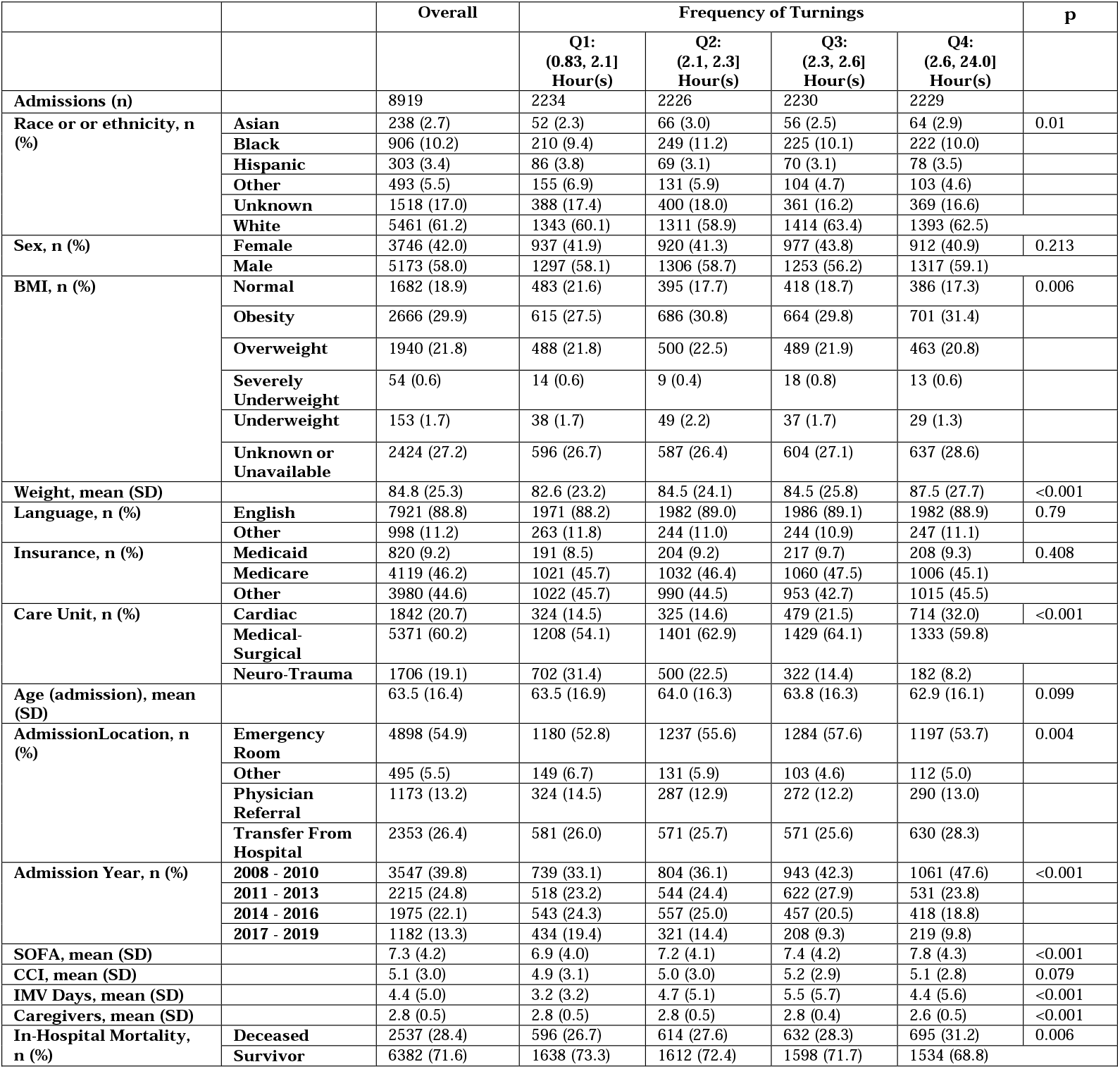
Cohort Description, stratified by frequency of turnings. BMI, body mass index; SOFA, sequential organ failure assessment score; CCI, Charlson comorbidity score; GCS, Glasgow coma score

**Table 1B.**
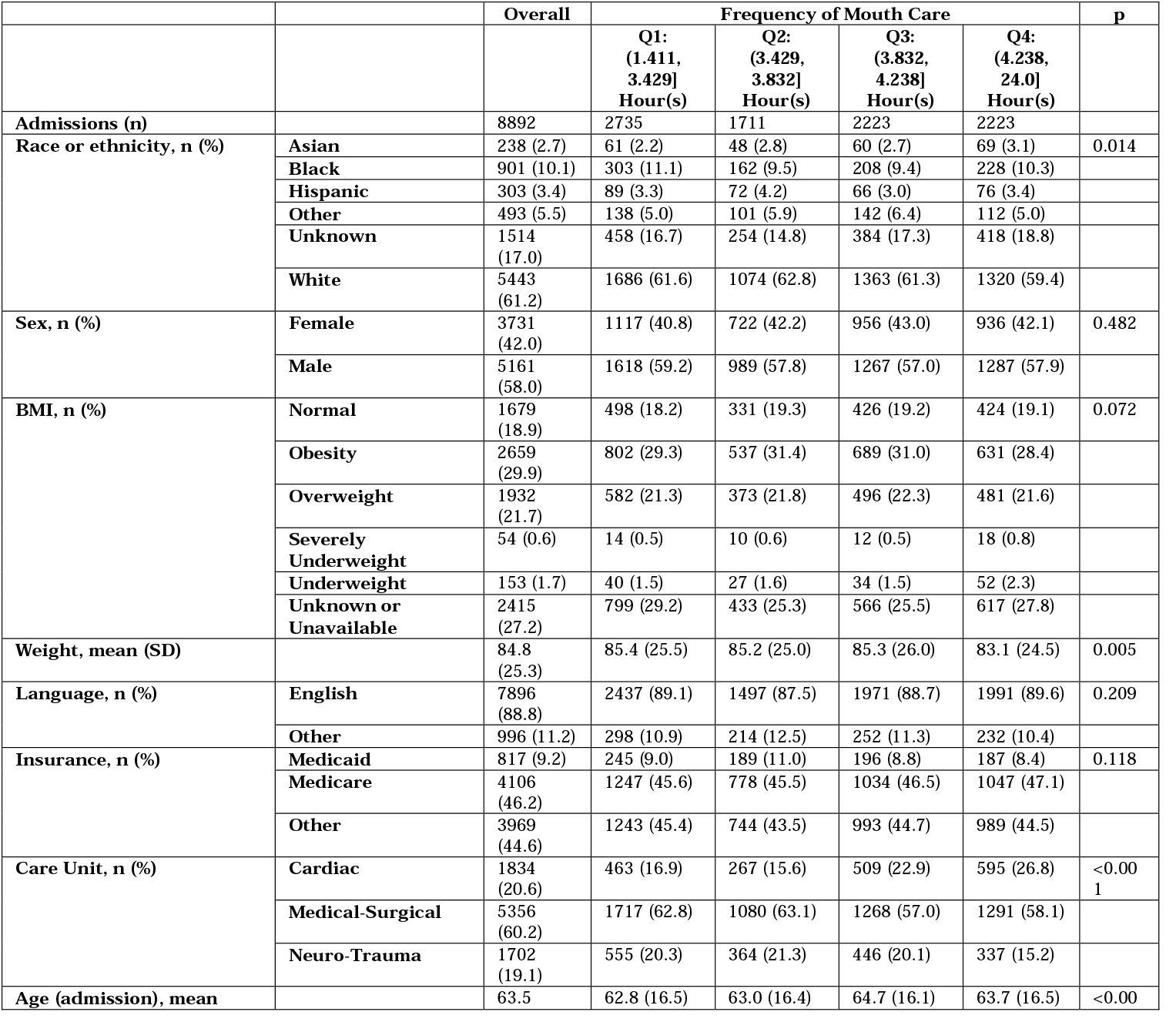
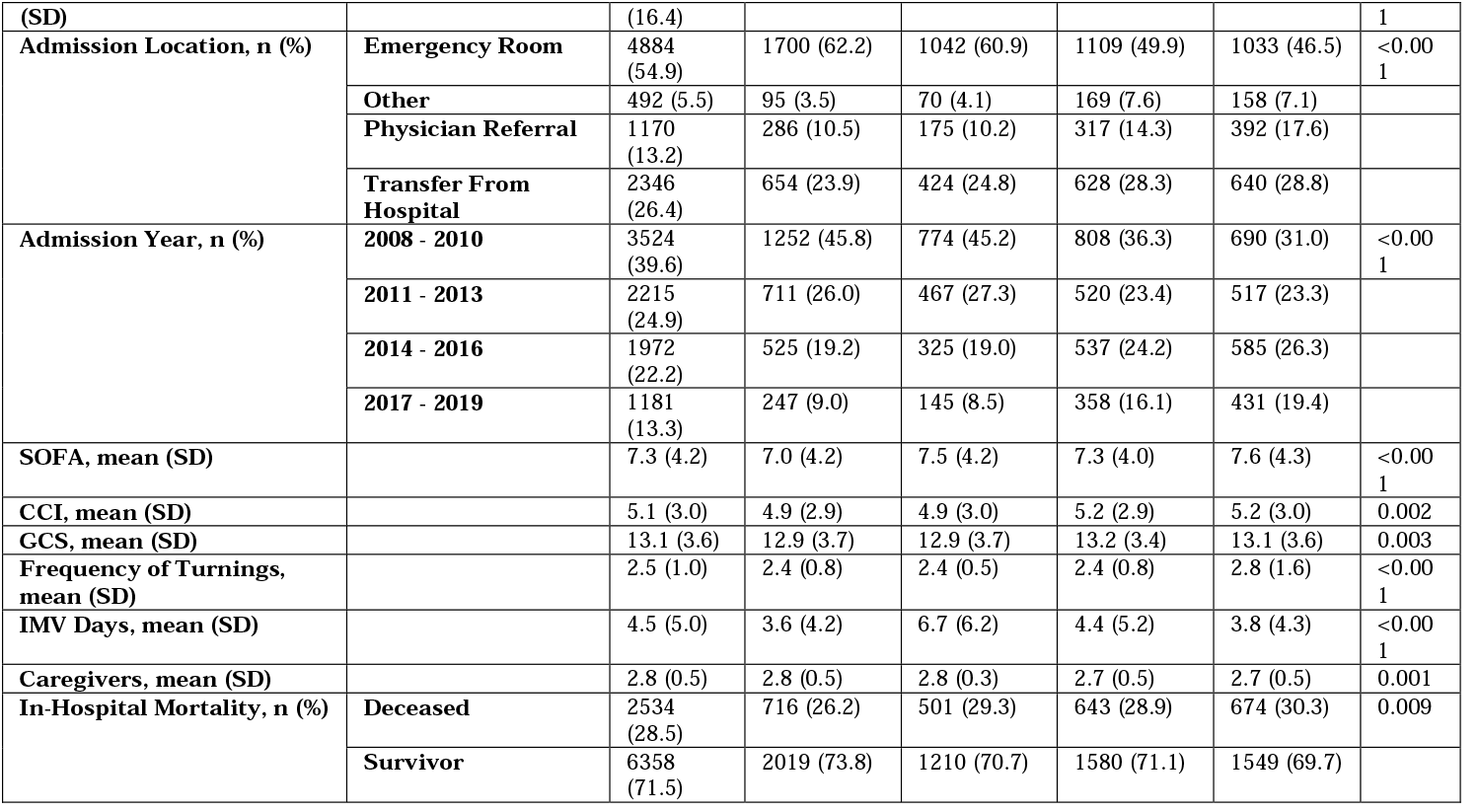
Cohort Description, stratified by frequency of mouth care. BMI, body mass index; SOFA, sequential organ failure assessment score; CCI, Charlson comorbidity score; GCS, Glasgow coma score

Our findings indicate that 50% of patients were turned significantly less than once every two hours (Median: 2.3 Hours, IQR: 2.1 - 2.6 Hours, p<<0.05), and that adherence to the protocol adherence was uneven - see figure 2. Focusing on weight as our variable of interest, we observe that heavier patients can expect to be turned less often, even compared to equivalently sick peers, in a predictable manner: for every extra weight decile, a patient can expect a reduction of one percentile in frequency of turning care (0.0760 turning interval percentile per weight percentile, p<<0.05, Figure 3a). More worryingly, patients who receive fewer turnings should also expect to receive fewer mouth care procedures, in a quantile ratio of 1 to 5 (0.2055 mouth care interval quantile per turning interval quantile, p<0.05, Figure 3b).

**Figure 2:**
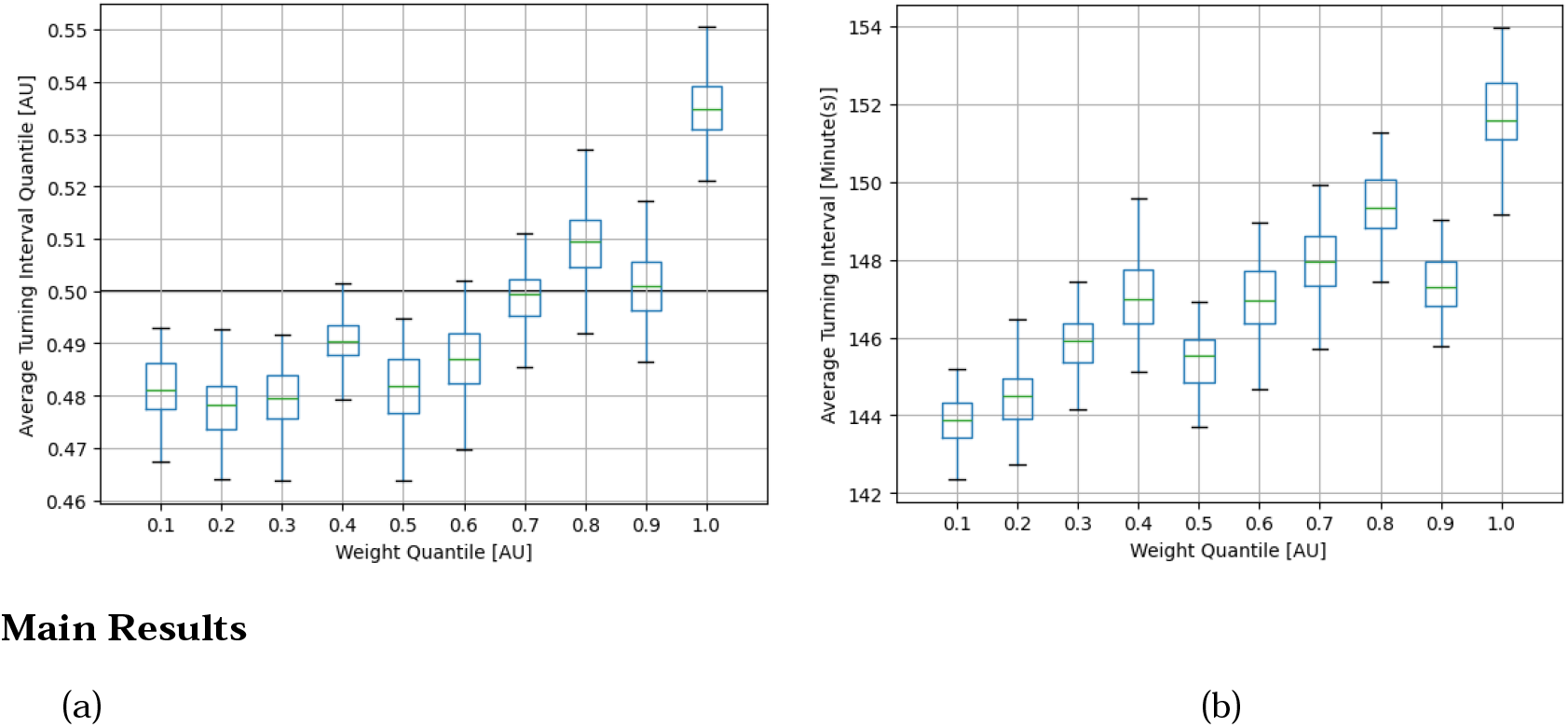
Bootstrap estimates of weight quantile against frequency of turnings (b) and frequency of turnings quantiles (a). (a) In a perfect scenario we would expect the behavior represented with the black line: all patients receive the same amount of care. In reality, we observe that patients on the right end of the spectrum in terms of weight quantiles can expect to receive turning care less frequently than those in weight quantiles towards the left. (b) Absolute average turning intervals. Patients are turned significantly less often than once every 120 minutes, with a marked skew towards heavier quantiles.

**Figure 3:**
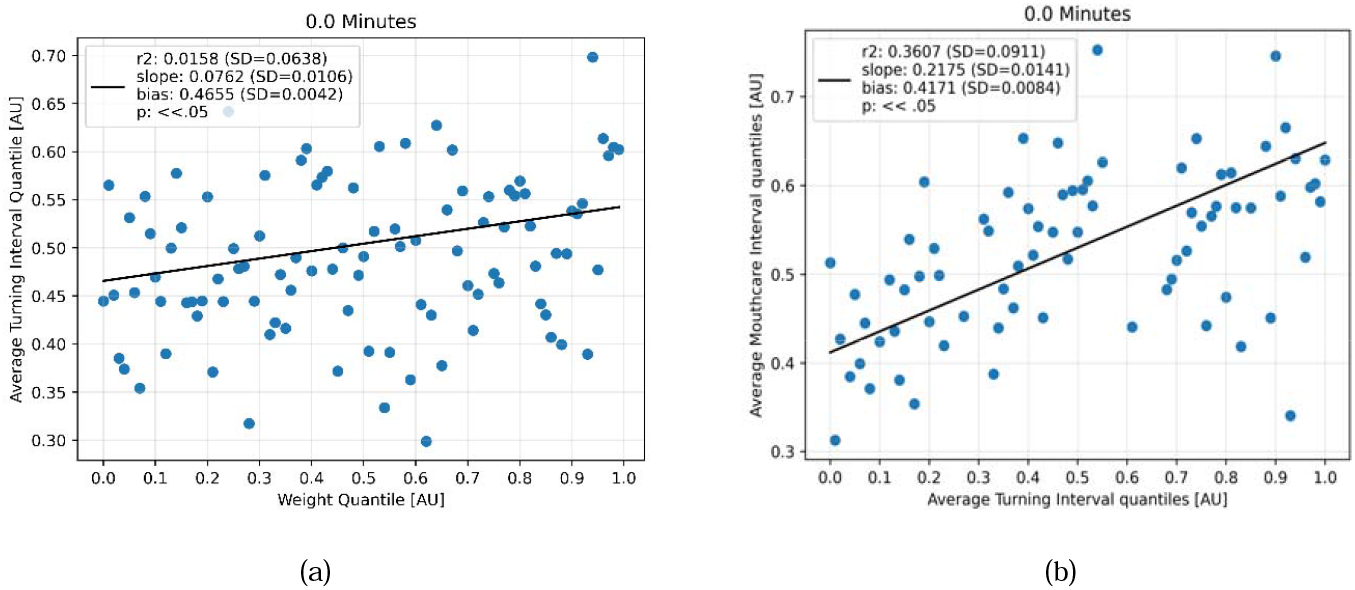
Univariate regression between weight quantiles and frequencies of turnings (a) and quantiles of turning and mouth care frequencies with no additive noise (b). There appears to be a significant positive correlation in both, suggesting that heavier patients who already receive fewer turns also receive less mouth care.

See Appendix A for an extensive description of the methodology and results.

## DISCUSSION

### Key Results

We investigated volumes of turnings and mouth care in a cohort of 8,919 Intensive Care Unit patients undergoing Invasive Mechanical Ventilation (IMV) identified within the MIMIC-IV database. Each patient was characterized by severity [20], comorbidity [21] and demography plus other non-clinical factors, namely ethnicity, type of health insurance and first spoken language. This resulted in a cohort with a majority of English speaking elderly white males (Mean Age: 63.5 years old at admission, 58% Males, 88.8% reported English as their first language).

In this cohort we observe that many IMV patients are turned significantly less often than required by standard protocols and the volume of care (measured by turning frequency) progressively diminishes as the patient’s weight increases. In addition, patients receiving less frequent turning also appeared to receive reduced mouth care. As such, our care phenotype here can be understood as those receiving suboptimal turning and mouthcare during invasive mechanical ventilation, with patient weight emerging as a key mediator of this observation. That worse care and reduced protocol adherence in one aspect (turning) then in turn spills over into another (mouth care) suggests a compounding effect of subtle diminishing care.

Besides the inherent practical importance of the specific finding itself, its implications are far-reaching. Our findings highlight the existence of a plane orthogonal to Social Determinants of Health (SDoH), intimately linked to the uniqueness of each site of care, operating on different axes: the phenotypes of care. These phenotypes directly profile the subtype of care a patient is actually receiving and complement our understanding of the SDoH, providing a tool for researchers using machine learning to move beyond the limitations of crude demographic labels to elicit new insights into care disparities.

This approach may be especially important for machine learning development. Often, SDoH or their proxies are naively integrated into models under the justification of fairness, but in reality, this may exacerbate biases. For instance, IMV turning prediction models trained on MIMIC might associate weight with ethnicity, systematically advising practitioners to turn heavier patients later, which is the opposite of what is intended, and this bias can also occur through ethnicity alone. The Care Phenotypes approach can thus support fairness evaluations for applied clinical algorithms.

Finally, our results indicate that no matter the amount of data or the sophistication of the machine learning models we employ, we will never resolve this issue unless we embed causal inference frameworks.[22] These frameworks should be designed in close collaboration with experienced clinical experts and should include a precise and attentive mapping of phenotypes of care derived from the unique circumstances in which the data was captured.

## Data Availability

The data produced in the present work are contained in the manuscript. Further information is available in appendices that will be added to the main manuscript.

## looking first at care received to create

- However, as ML approaches in clinical care are increasingly employed, it is clear that SDoH are not helpful for assessing if ML approaches are fair or if they increase inequality.
- In this paper, we argue that it is necessary to shift away from traditional demographic labels to more direct measures of quality of care received.
- We propose a novel approach, “Care Phenotypes” in order to assess the fairness of ML in relation to how Social Determinants of Health affect clinical care outcomes. The benefit of this method is that it directly measures the care that was received by patients, rather than relying on potentially inaccurate or complex demographic intersections.
- Care phenotypes speak to a growing body of concerns around how fairness in ML can be assessed and measured. It is important to clarify that Ca re phenotypes is not intended to be a direct intervention into clinical practice, rather it is a way to improve fairness evaluation for machine learning in healthcare. SDoC is an intervention that works in tandem with research on SDoH to improve fairness in clinical AI applications.
- This can be applied in fairness eval; causal inference work; disparity work.

